# Cardiovascular Complications of Modern Multiple Myeloma Therapy: Analysis of FDA Adverse Event Reporting System

**DOI:** 10.1101/2021.12.20.21268138

**Authors:** Zaki Al-Yafeai, Anil Ananthaneni, Mohamed Ghoweba, Hamzah Abduljabar, David Aziz, Udhayvir Grewal

**Author notes:** <u>Corresponding Author:</u> Zaki Al-Yafeai, MD, PhD, Internal Medicine Department, LSUHSC-Shreveport, Shreveport, LA. equal contribution (Anil Ananthaneni, Mohamed Ghoweba).

## Abstract

**Purpose:** Multiple myeloma accounts for over 15% of hematological malignancies. In attempt to tackle this malady, the FDA approved four drugs in 2015 which has propagated further development of new anti-multiple myeloma since. However, the health safety of these new agents is still ill-defined. The aim of this study is to delineate the cardiovascular adverse events of these drugs.

**Methods:** We searched the cardiac adverse events of the newly approved FDA drugs since 2015 using the U.S. Food and Drug Administration Adverse Events Reporting System database (FAERS). We calculated the reporting odds ratio (ROR) with 95% confidence for four drugs that have the highest incidence of cardiovascular adverse events.

**Results:** Among the medications that have approved for MM Between 2015-2020, four novel drugs showed the highest incidence of cardiotoxicity. ROR (95% CI) for atrial fibrillation due to elotuzumab, Ixazomib, daratumumab, and panobinostat compared to other FAERS drugs was 5.8 (4.4-7.7), 1.93 (1.5-2.3), 4.8 (4.2-5.6), and 5.7 (4.1-8.1), respectively. The ROR (95% CI) for cardiac failure was 8.2 (6.4-10.5), 4.7 (4.1-5.4), 5.8 (4.9-6.7), and 5.6 (3.8-8.1) and ROR (95% CI) for coronary disease was 2.7 (1.9-3.9), 2.7 (2.3-3.2), 2,3 (1.9-2.8), and 4.6 (3.2-6.6) due to elotuzumab, Ixazomib, daratumumab, and panobinostat versus all other drugs in FAERS.

**Conclusions:** The newly approved multiple myeloma therapy (elotuzumab, Ixazomib, daratumumab, panobinostat) are significantly associated with cardiotoxicity. These results highlight the importance of considering the cardiac history of patients with multiple myeloma when utilizing these novel agents.

## Introduction

Multiple myeloma is characterized by neoplastic proliferation of plasma cells that produce monoclonal immunoglobulins. Proliferation of cells in the bone marrow often cause osteolytic lesions, osteopenia and/or pathologic fractures, subsequent hypercalcemia stemming from bone destruction and anemia mainly due to bone marrow replacement (Oyajobi, 2007). There is also observed renal dysfunction due to accumulation and precipitation of light chains which form casts in distal renal tubules (Dimopoulos, Kastritis, Rosinol, Blade, & Ludwig, 2008). It is estimated that 35000 new cases are diagnosed annually, accounting for nearly 2% of all new cancer diagnoses^3^. Age-adjusted death rates have not changed significantly over 2009–2018 and have remained at around 3.1 per 100,000 population(Ludwig, Novis Durie, Meckl, Hinke, & Durie, 2020).

While the initial therapy before newer therapeutic advances has classically consisted of VRd (bortezomib + lenalinomide + dexamethasone) and/or stem cell transplant with bortezomib/lenalinomide maintenance(Kumar et al., 2017), we have had newer approvals for treatment of multiple myeloma which have received at least one previous regimen. Between 2015-2016, the FDA approved 4 new drugs – elotuzumab (activates NK cells)(Magen & Muchtar, 2016), ixazomib (proteosome inhibitor)(Raedler, 2016), daratumumab (targeting CD38)(Sanchez, Wang, Siegel, & Wang, 2016), panobinostat(histone deacetylase inhibitor)(Laubach, Moreau, San-Miguel, & Richardson, 2015) and more recently belantamab and selinexor in 2020(Joseph, Tai, Anderson, & Lonial, 2021; Offidani, Corvatta, More, & Olivieri, 2021). Clinical Daratumumab is frequently used in first line regimens along with VRd as D-VRd(Kumar et al., 2017).

While the usage of these drugs has increased in the recent years, there is little published evidence about their cardiovascular adverse effect profile. One of the largest trials, the CASTOR trial that included 283 and 286 patients in the VRd and D-VRd arms respectively reported negligible cardiovascular side effects – 2 patients with atrial fibrillation in both arms, 1 patient with acute coronary syndrome among others in the D-VRd arm that could not reach statistical significance given low sample size(Palumbo et al., 2016). Therefore, we sought to analyze data from FAERS to study the reported cardiovascular adverse events of the newly approved multiple myeloma.

## Methods

This is a retrospective adverse event analysis of the U.S. Food and Drug Administration’s Adverse Event Reporting System (FEARS) database. FAERS is an online publicly available post-marketing pharmacovigilance database that records millions of adverse events, medication error reports, and product quality complaints submitted by healthcare professionals, manufacturers, and consumers worldwide for approved drugs and biologic products. Reports are evaluated by clinical reviewers in the Center for Drug Evaluation and Research and the Center for Biologics Evaluation and Research. The database includes data regarding the suspected pharmaceutical agent, indication for its use, patient characteristics, as well as the reported adverse event including its date of occurrence, nature, and outcome, among others. Since the data is anonymized, approval by an ethics committee was not required for this analysis. We aimed to evaluate the cardiovascular adverse events associated with newly FDA-approved multiple-myeloma therapeutic agents. Queries for the specific adverse event terms were performed (Supplemental). Statistical testing to measure the association of these drugs with the specified cardiac events of interest was performed using disproportionality signal analysis. These were presented as reporting odds ratios (RORs) of reported cardiac adverse events concomitant with the use of a newly approved multiple myeloma agent compared to the same reported adverse events within the entire database of pharmaceuticals within FAERS. The ROR was considered significant when the lower limit of the 95% CI was >1.0.

## Results

The total number of adverse events (AEs) from all drugs in the FAERS database was over 20 million. Of those, the total number of AEs caused by the newly-approved therapeutic agents for multiple myeloma (including in combination with other medications) evaluated in this study was 30,797. Ixazomib had the highest number of total reported AEs with 13,701, followed by daratumumab with 10,235, while belantamab mafodotin had the lowest with 351 AEs. Out of the 30,797 AEs reported, 1131 were cardiac AEs. These included heart failure, atrial fibrillation, and coronary heart disease. Heart failure was reported the most with 470 AEs, while atrial fibrillation came next with 346, and coronary heart disease with 315. Ixazomib had the highest reported heart failure and coronary heart disease AEs with 189 and 145 AEs respectively. Daratumumab had the highest atrial fibrillation AEs at 164 [Table 2].

Table 1 and supplemental tables 1-6 further demonstrate the characteristics of the cardiac related AEs associated with Elotuzumab, Ixazomib, Daratumumab, Panobinostat, Selinexor, and Belantamab mafodotin. As shown in these tables, male predominance was seen amongst all the drug-related AEs reported. For all the drugs, at least 98% of the reported AEs were classified as serious, with the mortality ranging from 19%-46% amongst these serious outcomes.

Table 1
|  | Atrial Fibrillation | Cardiac Failure | Coronary Artery Disease |
| --- | --- | --- | --- |
| **Elotuzumab** |  |  |  |
| Total number of reported AEs | 50 | 61 | 26 |
| **Age in years** |  |  |  |
| 18-64 | 14 | 14 | 7 |
| 65-85 | 30 | 39 | 13 |
| 85+ | 0 | 1 | 0 |
| Not Specified | 6 | 7 | 6 |
| **Outcome reported per AE** |  |  |  |
| Hospitalization | 39 | 40 | 14 |
| Life-threatening | 6 | 4 | 7 |
| Death | 11 | 13 | 16 |
| Other Outcome | 40 | 52 | 16 |
| Disabled | 1 | 4 | 0 |
| **Ixazomib** |  |  |  |
| Total number of reported AEs | 93 | 212 | 139 |
| **Age in years** |  |  |  |
| 18-64 | 10 | 21 | 21 |
| 65-85 | 61 | 125 | 77 |
| 85+ | 3 | 16 | 5 |
| Not Specified | 19 | 50 | 36 |
| **Outcome reported per AE** |  |  |  |
| Hospitalization | 72 | 145 | 88 |
| Life-threatening | 5 | 14 | 34 |
| Nonserious | 1 | 2 | 0 |
| Death | 18 | 50 | 46 |
| Other Outcome | 48 | 179 | 112 |
| Disabled | 0 | 4 | 1 |
| **Daratumumab** |  |  |  |
| Total number of reported AEs | 191 | 207 | 94 |
| **Age in years** |  |  |  |
| 18-64 | 24 | 31 | 19 |
| 65-85 | 115 | 99 | 52 |
| 85+ | 2 | 10 | 3 |
| Not Specified | 50 | 67 | 20 |
| **Outcome reported per AE** |  |  |  |
| Hospitalization | 116 | 134 | 70 |
| Life-threatening | 12 | 35 | 24 |
| Death | 36 | 69 | 29 |
| Other Outcome | 134 | 139 | 37 |
| Disabled | 1 | 4 | 4 |
| **Panobinostat** |  |  |  |
| Total number of reported AEs | 33 | 32 | 19 |
| **Age in years** |  |  |  |
| 7-12 | 0 | 1 | 0 |
| 18-64 | 3 | 5 | 7 |
| 65-85 | 16 | 14 | 3 |
| 85+ | 0 | 1 | 0 |
| Not Specified | 14 | 11 | 9 |
| **Outcome reported per AE** |  |  |  |
| Hospitalization | 4 | 3 | 4 |
| Life-threatening | 1 | 1 | 1 |
| Nonserious | 0 | 0 | 0 |
| Death | 1 | 1 | 1 |
| Other Outcome | 3 | 4 | 5 |

able 2
| Drug Name | Total<br>(with<br>others+<br>Alone) | Heart<br>Failure | Atrial<br>Fibrillation | Coronary<br>Heart<br>Disease | Total<br>Cardiac<br>Related AEs | Total<br>Adverse<br>Events |
| --- | --- | --- | --- | --- | --- | --- |
| Elotuzumab | Total | 59 (2.3%) | 49 (2%) | 27(1%) | 135 | 2511 |
|  | Alone | 10 | 7 | 1 | 18 |  |
| Ixazomib | Total | 189 (7.5%) | 89 (3.5%) | 145 (5.8%) | 423 | 13701 |
|  | Alone | 48 | 12 | 27 | 87 |  |
| Daratumumab | Total | 172 (6.85%) | 164(6.5%) | 107(4.3%) | 443 | 10235 |
|  | Alone | 52 | 36 | 31 | 119 |  |
| Panobinostat | Total | 28 (1.1%) | 33 (1.3%) | 21 (0.8%) | 82 | 1721 |
|  | Alone | 6 | 7 | 5 | 18 |  |
| Belantamab<br>mafodotin | Total | 3 (0.1%) | 5 (0.2%) | 6 (0.2%) | 14 | 351 |
|  | Alone | 3 | 3 | 5 | 11 |  |
| Selinexor | Total | 19 (0.76%) | 6 (0.24%) | 9 (0.36%) | 34 | 2278 |
|  | Alone | 8 | 3 | 6 | 17 |  |

According to disproportionality signal analysis and compared to other medications, atrial fibrillation and coronary heart disease were mostly associated with the use of panobinostat with ROR of 5.7 (95% CI: 4.1-8.1, P<0.0001) and 4.6 (95% CI: 3.8-8.1, P<0.0001) and elotuzumab with ROR of 5.8 (95% CI: 4.4-7.7, P<0.0001) and 2.7 (95% CI: 1.9-3.9, P<0.0001) respectively. Coronary artery disease had the lowest ROR with all agents with odds ratio between 2.3-4.6. Heart failure was reported the most with the use of elotuzumab with ROR of 8.2 (95% CI: 6.4-10.5, P<0.0001), while ixazomib exhibited the least association with ROR of 4.7 (95% CI: 4.1-5.4, P<0.0001). Of note, panobinostat’s and daratumumab’s ROR values were closely related compared to other agents [Figure 1 A-C and Table 3].

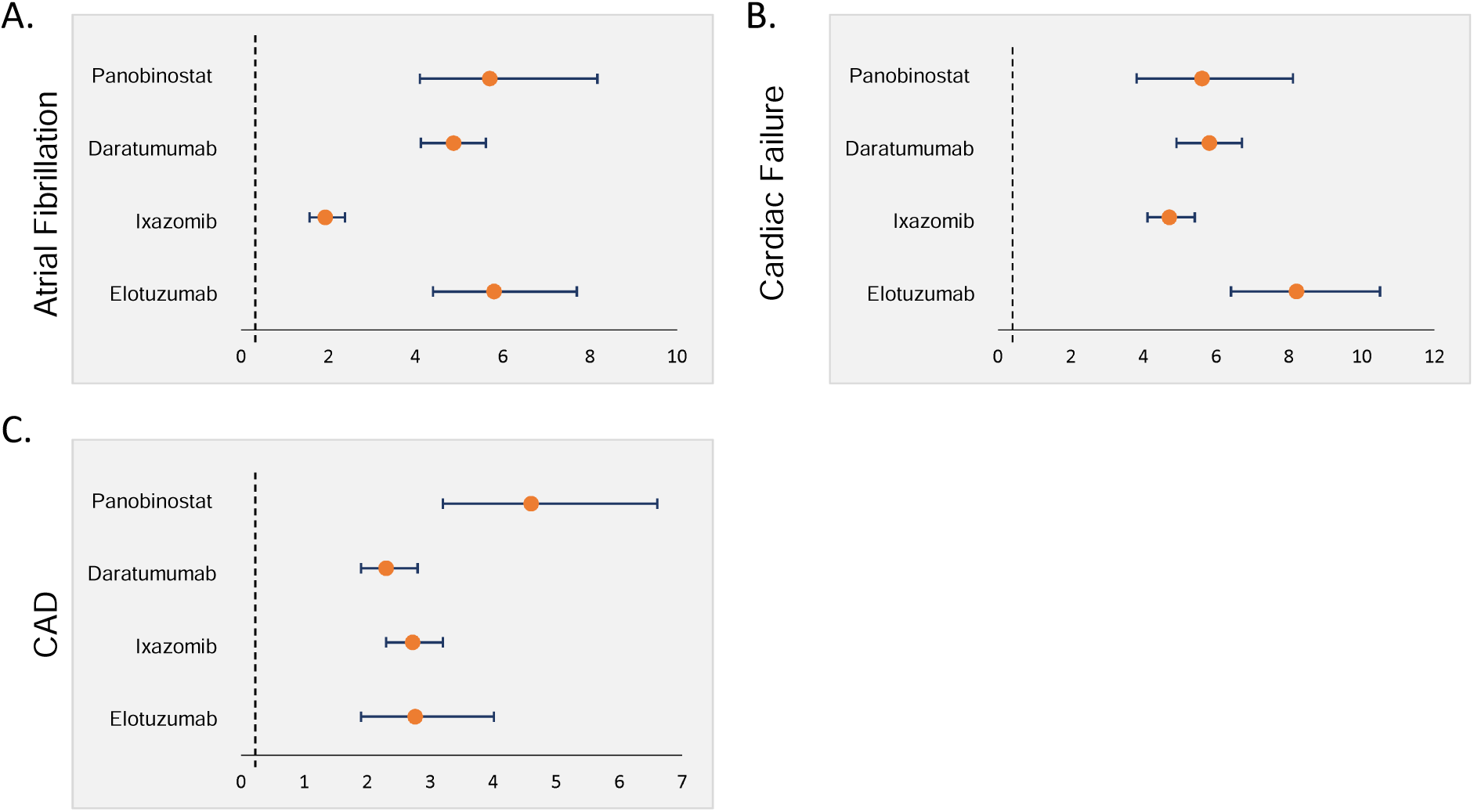

Table 3
| Drug name | Elotuzumab vs others | Ixazomib and vs others | Daratumumab vs others | Panobinostat vs others |
| --- | --- | --- | --- | --- |
| **Atrial Fibrillation** | 5.84 (4.4-7.7)<br>P<0.0001 | 1.93 (1.55-2.36)<br>P<0.0001 | 4.87 (4.2-5.6)<br>P<0.0001 | 5.74 (4.1-8.1)<br>P<0.0001 |
| **Cardiac failure** | 8.2 (6.4-10.5)<br>P<0.0001 | 4.7 (4.1-5.4)<br>P<0.0001 | 5.8 (4.9-6.7)<br>P<0.0001 | 5.6 (3.8-8.1)<br>P<0.0001 |
| **CAD** | 2.76 (1.9-3.9)<br>P<0.0001 | 2.7 (2.3-3.2)<br>P<0.0001 | 2.3 (1.9-2.8)<br>P<0.0001 | 4.6 (3.2-6.6)<br>P<0.0001 |

## Discussion

The results of this study demonstrated an association between the use of the newest novel multiple myeloma agents and cardiotoxicity. Among all of the adverse events reported for these new agents on FAERS, cardiac complications represented little less than 10%. Of all of the newly FDA approved multiple myeloma agents, elotuzumab, Ixazomib, daratumumab, panobinostat showed significant cardiac adverse events. Interestingly, cardiac failure, atrial fibrillation and coronary artery diseases were profoundly the highest of all reported cardiotoxicity. Interestingly Isatuximab, and belantamab mafodotin showed safer cardiac profile compared to elotuzumab, Ixazomib, daratumumab, panobinostat.

The fascinating Eloquent trials showed that elotuzumab significantly reduced multiple myeloma progression. Unspecified cardiac adverse events were reported among 7 patients with one death from heart failure with no concerning effects on QT prolongation(Dimopoulos et al., 2018; Passey, Darbenzio, Jou, Lynch, & Gupta, 2016). The TOURMALINE trials that demonstrated the efficacy of Ixazomib on multiple myeloma reported similar adverse cardiac events (heart failure, myocardial infarction and arrhythmia) between the Ixazomib and the placebo group(Dimopoulos et al., 2020; Facon et al., 2021; Moreau et al., 2016). Three cases of myocardial infarction reported with panobinostat compared to placebo group(San-Miguel et al., 2014). Additionally,17.6% of patient who received panobinostat and bortezomib and dexamethasone reported cardiac complications (most frequently atrial fibrillation, tachycardia, palpitation, and sinus tachycardia) on phase 3 PANORAMA-1 trial(Plummer, Driessen, Szabo, & Mateos, 2019). Interestingly, no major cardiac adverse events reported associated with daratumumab(Dimopoulos et al., 2021; Palumbo et al., 2016).

While the findings of this study are novel, certain limitations pertains to this study. The wholistic profile of patient (comorbidities, risk factors, family history…etc.) are unknown. Moreover, prior cardiovascular functioning and other cardiac disorders are not considered when performing these studies. Additionally, the fundamental mechanisms mediating the cardiotoxicity associated with the novel multiple myeloma agents remain elusive.

The significance of our data stems from the increasing use of the new agents for multiple myeloma. As addressed above, multiple clinical trials showed sporadic cardiac adversaries of the novel multiple myeloma agents, however, a more thorough investigation of the cardiovascular profile of these medications is lacking. Our study strongly agrees with the aforementioned studies and showed enhanced cardiotoxicity especially atrial fibrillation, coronary artery disease and heart failure. Based on our results, we recommend physicians to discuss the cardiovascular risk with patients upon starting these medications. Electrocardiography and echocardiogram maybe help establish baseline cardiac function prior initiating these regimens.

In conclusions, the novel multiple myeloma agents are associated with enhance atrial fibrillation, coronary artery disease and heart failure. Patient risk stratification and baseline cardiac functioning testing (electrocardiography and echocardiogram) should be considered upon initiating these agents.

## Data Availability

All data produced in the present study are available upon reasonable request to the authors

## Acknowledgment

None.

## Statements and Declarations

### Author Contributions

All the authors contributed to the study design, writing and analysis. Data collection and analysis [Zaki Al-Yafeai, Anil Ananthaneni, Mohamed Ghoweba], design [ Zaki Al-Yafeai, Hamzah Abduljabar, David Aziz, and Udhayvir Grewal]. The first draft was written by [Zaki Al-Yafeai] with edits and revision from all the authors. All authors read and approved the manuscript.

### Data Availability

The datasets from the current study are available from the corresponding author upon request

