## Supplementary material for "Cardiovascular Complications of Modern Multiple Myeloma Therapy: Analysis of FDA Adverse Event Reporting System": Supp

### Slide 1
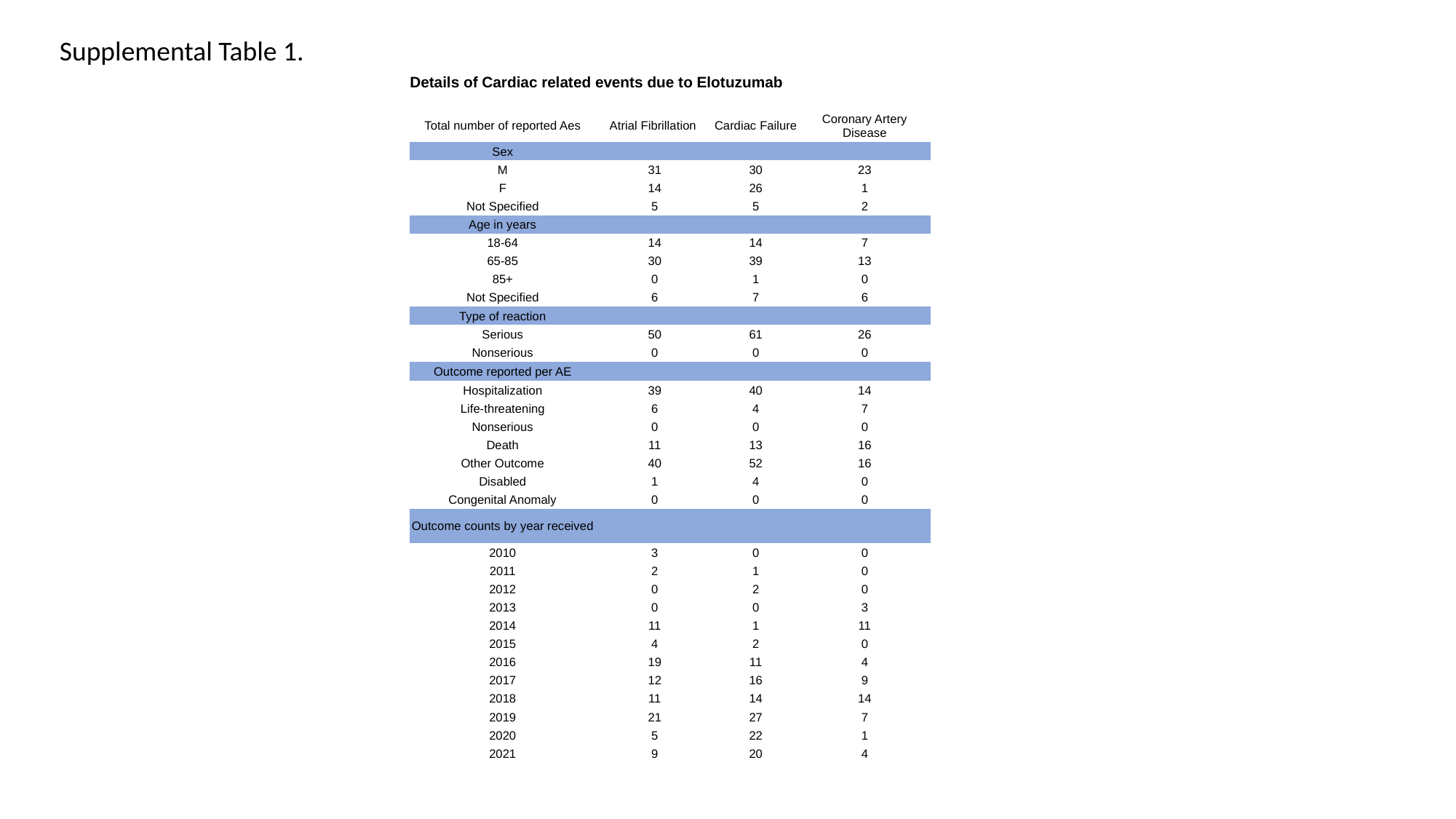

Supplemental Table 1.
| Details of Cardiac related events due to Elotuzumab | | | |
| --- | --- | --- | --- |
| Total number of reported Aes | Atrial Fibrillation | Cardiac Failure | Coronary Artery Disease |
| Sex | | | |
| M | 31 | 30 | 23 |
| F | 14 | 26 | 1 |
| Not Specified | 5 | 5 | 2 |
| Age in years | | | |
| 18-64 | 14 | 14 | 7 |
| 65-85 | 30 | 39 | 13 |
| 85+ | 0 | 1 | 0 |
| Not Specified | 6 | 7 | 6 |
| Type of reaction | | | |
| Serious | 50 | 61 | 26 |
| Nonserious | 0 | 0 | 0 |
| Outcome reported per AE | | | |
| Hospitalization | 39 | 40 | 14 |
| Life-threatening | 6 | 4 | 7 |
| Nonserious | 0 | 0 | 0 |
| Death | 11 | 13 | 16 |
| Other Outcome | 40 | 52 | 16 |
| Disabled | 1 | 4 | 0 |
| Congenital Anomaly | 0 | 0 | 0 |
| Outcome counts by year received | | | |
| 2010 | 3 | 0 | 0 |
| 2011 | 2 | 1 | 0 |
| 2012 | 0 | 2 | 0 |
| 2013 | 0 | 0 | 3 |
| 2014 | 11 | 1 | 11 |
| 2015 | 4 | 2 | 0 |
| 2016 | 19 | 11 | 4 |
| 2017 | 12 | 16 | 9 |
| 2018 | 11 | 14 | 14 |
| 2019 | 21 | 27 | 7 |
| 2020 | 5 | 22 | 1 |
| 2021 | 9 | 20 | 4 |

### Slide 2
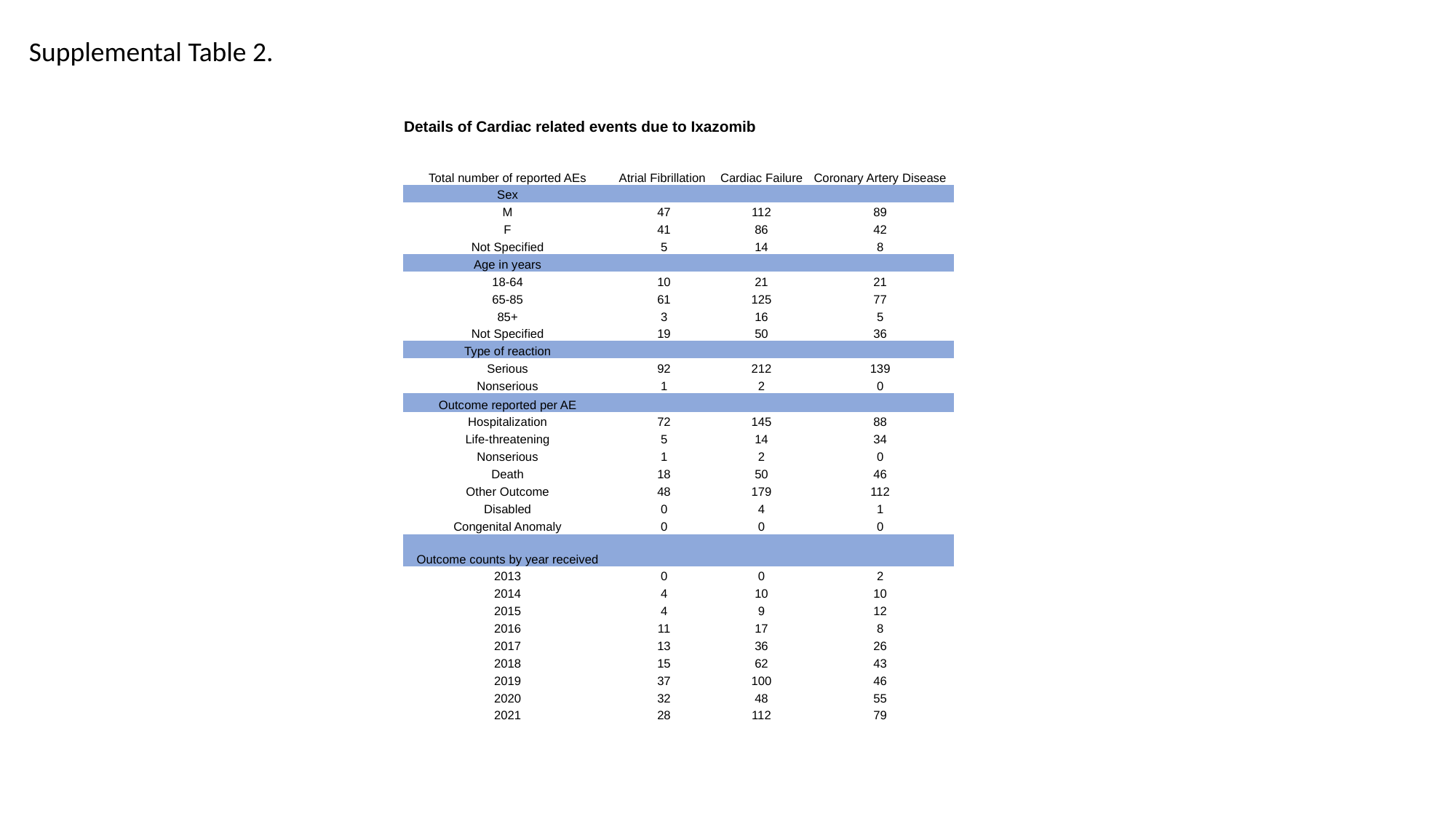

Supplemental Table 2.
| Details of Cardiac related events due to Ixazomib | | | |
| --- | --- | --- | --- |
| Total number of reported AEs | Atrial Fibrillation | Cardiac Failure | Coronary Artery Disease |
| Sex | | | |
| M | 47 | 112 | 89 |
| F | 41 | 86 | 42 |
| Not Specified | 5 | 14 | 8 |
| Age in years | | | |
| 18-64 | 10 | 21 | 21 |
| 65-85 | 61 | 125 | 77 |
| 85+ | 3 | 16 | 5 |
| Not Specified | 19 | 50 | 36 |
| Type of reaction | | | |
| Serious | 92 | 212 | 139 |
| Nonserious | 1 | 2 | 0 |
| Outcome reported per AE | | | |
| Hospitalization | 72 | 145 | 88 |
| Life-threatening | 5 | 14 | 34 |
| Nonserious | 1 | 2 | 0 |
| Death | 18 | 50 | 46 |
| Other Outcome | 48 | 179 | 112 |
| Disabled | 0 | 4 | 1 |
| Congenital Anomaly | 0 | 0 | 0 |
| Outcome counts by year received | | | |
| 2013 | 0 | 0 | 2 |
| 2014 | 4 | 10 | 10 |
| 2015 | 4 | 9 | 12 |
| 2016 | 11 | 17 | 8 |
| 2017 | 13 | 36 | 26 |
| 2018 | 15 | 62 | 43 |
| 2019 | 37 | 100 | 46 |
| 2020 | 32 | 48 | 55 |
| 2021 | 28 | 112 | 79 |

### Slide 3
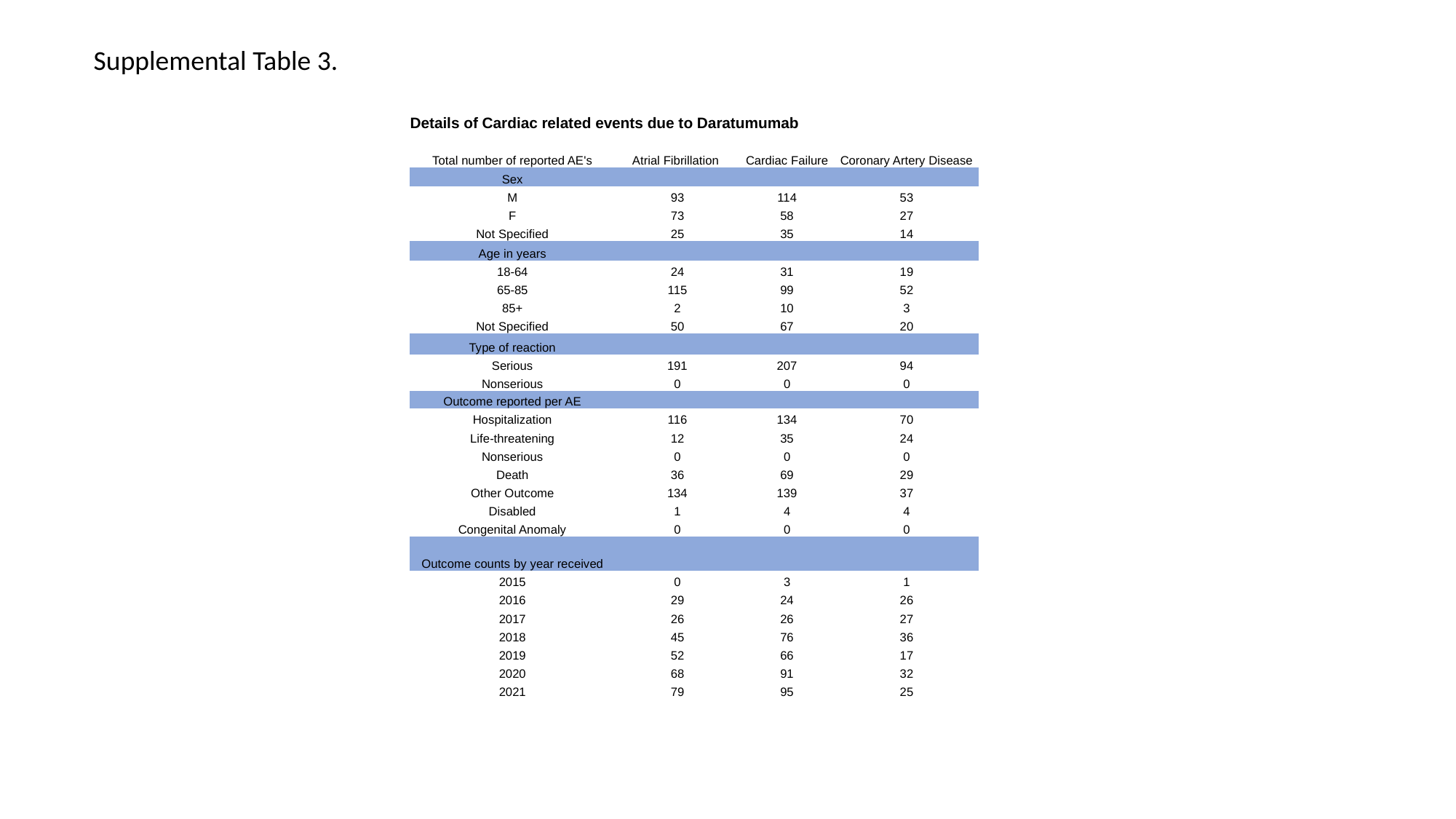

Supplemental Table 3.
| Details of Cardiac related events due to Daratumumab | | | |
| --- | --- | --- | --- |
| Total number of reported AE's | Atrial Fibrillation | Cardiac Failure | Coronary Artery Disease |
| Sex | | | |
| M | 93 | 114 | 53 |
| F | 73 | 58 | 27 |
| Not Specified | 25 | 35 | 14 |
| Age in years | | | |
| 18-64 | 24 | 31 | 19 |
| 65-85 | 115 | 99 | 52 |
| 85+ | 2 | 10 | 3 |
| Not Specified | 50 | 67 | 20 |
| Type of reaction | | | |
| Serious | 191 | 207 | 94 |
| Nonserious | 0 | 0 | 0 |
| Outcome reported per AE | | | |
| Hospitalization | 116 | 134 | 70 |
| Life-threatening | 12 | 35 | 24 |
| Nonserious | 0 | 0 | 0 |
| Death | 36 | 69 | 29 |
| Other Outcome | 134 | 139 | 37 |
| Disabled | 1 | 4 | 4 |
| Congenital Anomaly | 0 | 0 | 0 |
| Outcome counts by year received | | | |
| 2015 | 0 | 3 | 1 |
| 2016 | 29 | 24 | 26 |
| 2017 | 26 | 26 | 27 |
| 2018 | 45 | 76 | 36 |
| 2019 | 52 | 66 | 17 |
| 2020 | 68 | 91 | 32 |
| 2021 | 79 | 95 | 25 |

### Slide 4
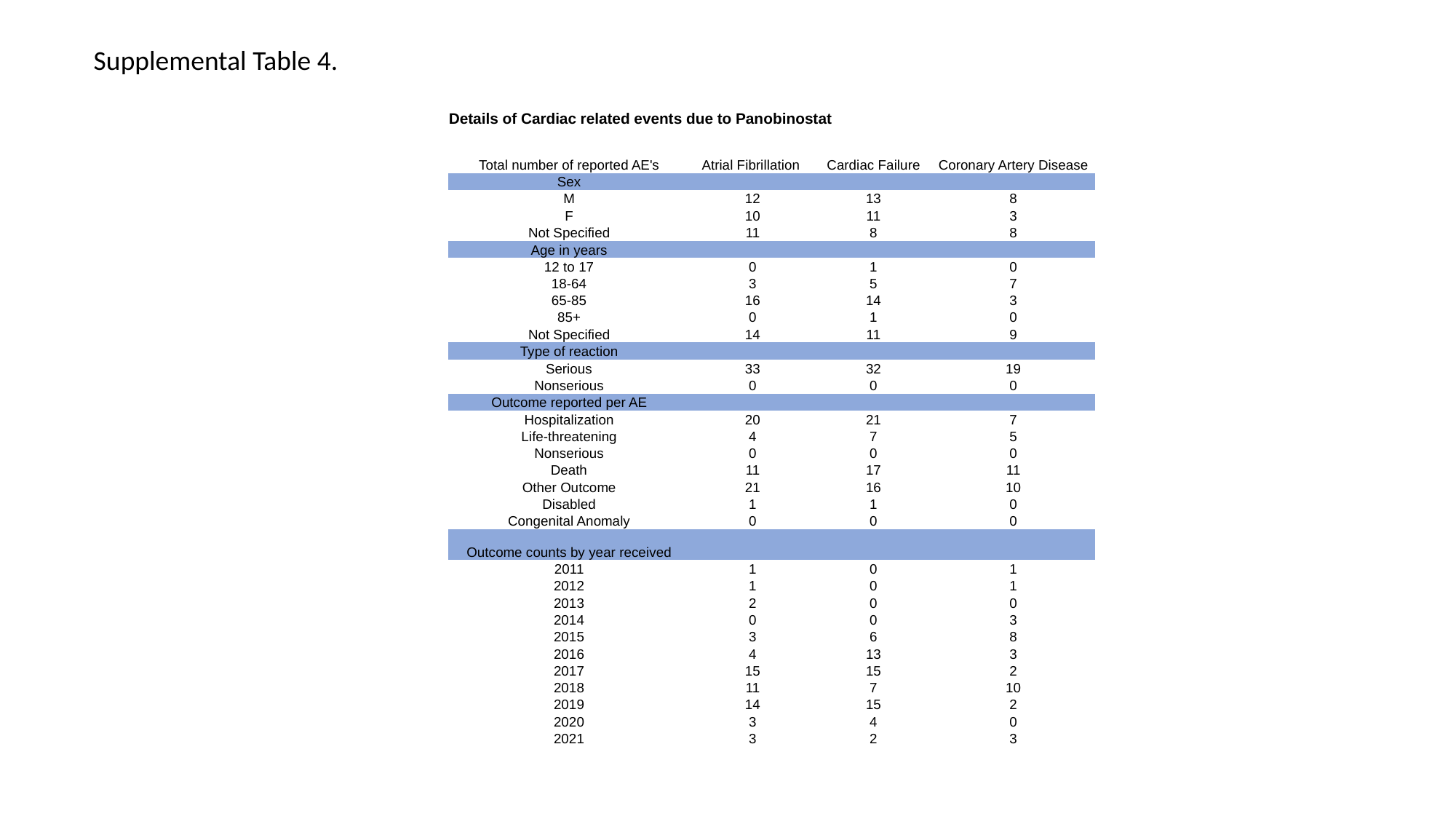

Supplemental Table 4.
| Details of Cardiac related events due to Panobinostat | | | |
| --- | --- | --- | --- |
| Total number of reported AE's | Atrial Fibrillation | Cardiac Failure | Coronary Artery Disease |
| Sex | | | |
| M | 12 | 13 | 8 |
| F | 10 | 11 | 3 |
| Not Specified | 11 | 8 | 8 |
| Age in years | | | |
| 12 to 17 | 0 | 1 | 0 |
| 18-64 | 3 | 5 | 7 |
| 65-85 | 16 | 14 | 3 |
| 85+ | 0 | 1 | 0 |
| Not Specified | 14 | 11 | 9 |
| Type of reaction | | | |
| Serious | 33 | 32 | 19 |
| Nonserious | 0 | 0 | 0 |
| Outcome reported per AE | | | |
| Hospitalization | 20 | 21 | 7 |
| Life-threatening | 4 | 7 | 5 |
| Nonserious | 0 | 0 | 0 |
| Death | 11 | 17 | 11 |
| Other Outcome | 21 | 16 | 10 |
| Disabled | 1 | 1 | 0 |
| Congenital Anomaly | 0 | 0 | 0 |
| Outcome counts by year received | | | |
| 2011 | 1 | 0 | 1 |
| 2012 | 1 | 0 | 1 |
| 2013 | 2 | 0 | 0 |
| 2014 | 0 | 0 | 3 |
| 2015 | 3 | 6 | 8 |
| 2016 | 4 | 13 | 3 |
| 2017 | 15 | 15 | 2 |
| 2018 | 11 | 7 | 10 |
| 2019 | 14 | 15 | 2 |
| 2020 | 3 | 4 | 0 |
| 2021 | 3 | 2 | 3 |

### Slide 5
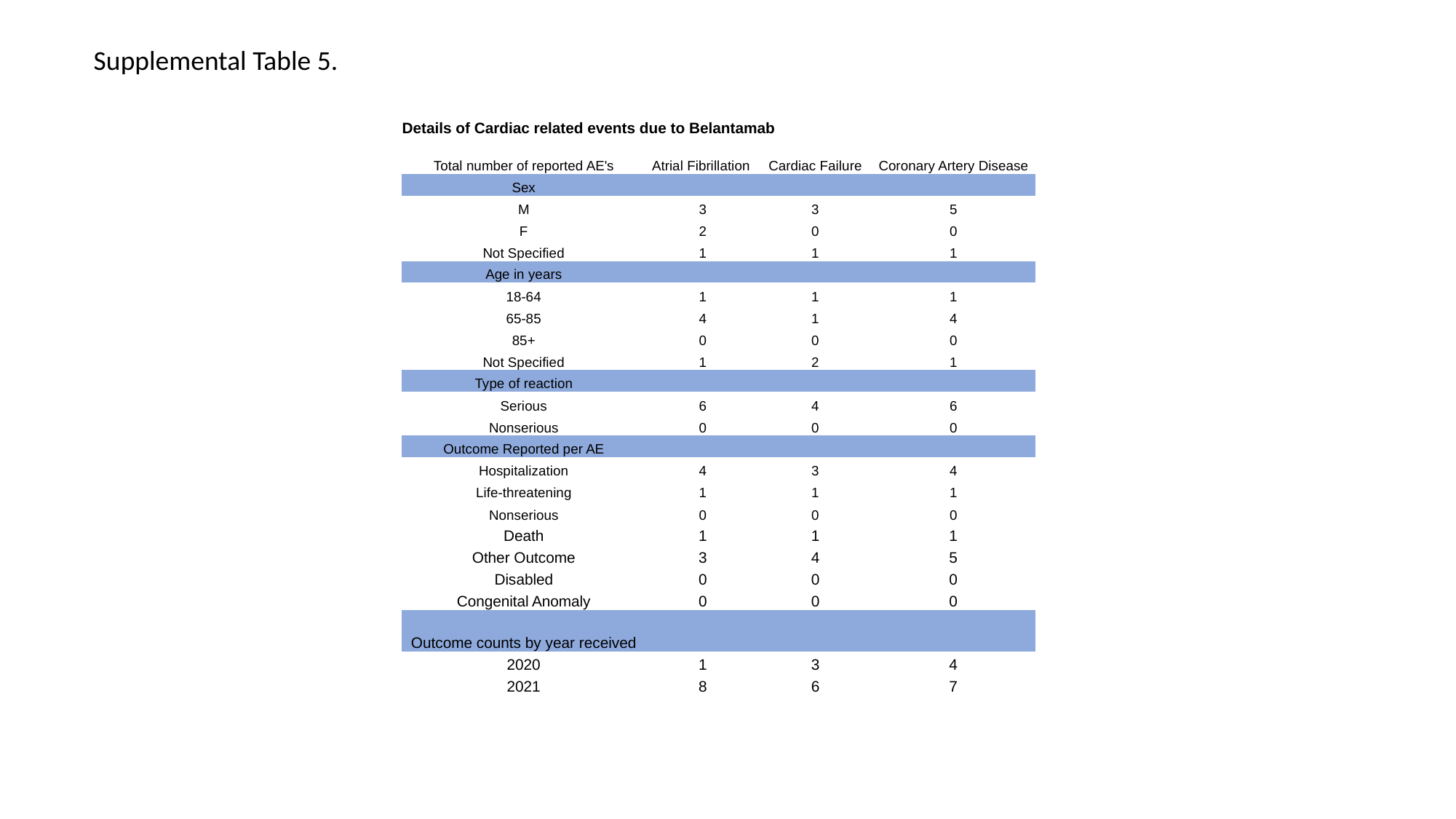

Supplemental Table 5.
| Details of Cardiac related events due to Belantamab | | | |
| --- | --- | --- | --- |
| Total number of reported AE's | Atrial Fibrillation | Cardiac Failure | Coronary Artery Disease |
| Sex | | | |
| M | 3 | 3 | 5 |
| F | 2 | 0 | 0 |
| Not Specified | 1 | 1 | 1 |
| Age in years | | | |
| 18-64 | 1 | 1 | 1 |
| 65-85 | 4 | 1 | 4 |
| 85+ | 0 | 0 | 0 |
| Not Specified | 1 | 2 | 1 |
| Type of reaction | | | |
| Serious | 6 | 4 | 6 |
| Nonserious | 0 | 0 | 0 |
| Outcome Reported per AE | | | |
| Hospitalization | 4 | 3 | 4 |
| Life-threatening | 1 | 1 | 1 |
| Nonserious | 0 | 0 | 0 |
| Death | 1 | 1 | 1 |
| Other Outcome | 3 | 4 | 5 |
| Disabled | 0 | 0 | 0 |
| Congenital Anomaly | 0 | 0 | 0 |
| Outcome counts by year received | | | |
| 2020 | 1 | 3 | 4 |
| 2021 | 8 | 6 | 7 |

### Slide 6
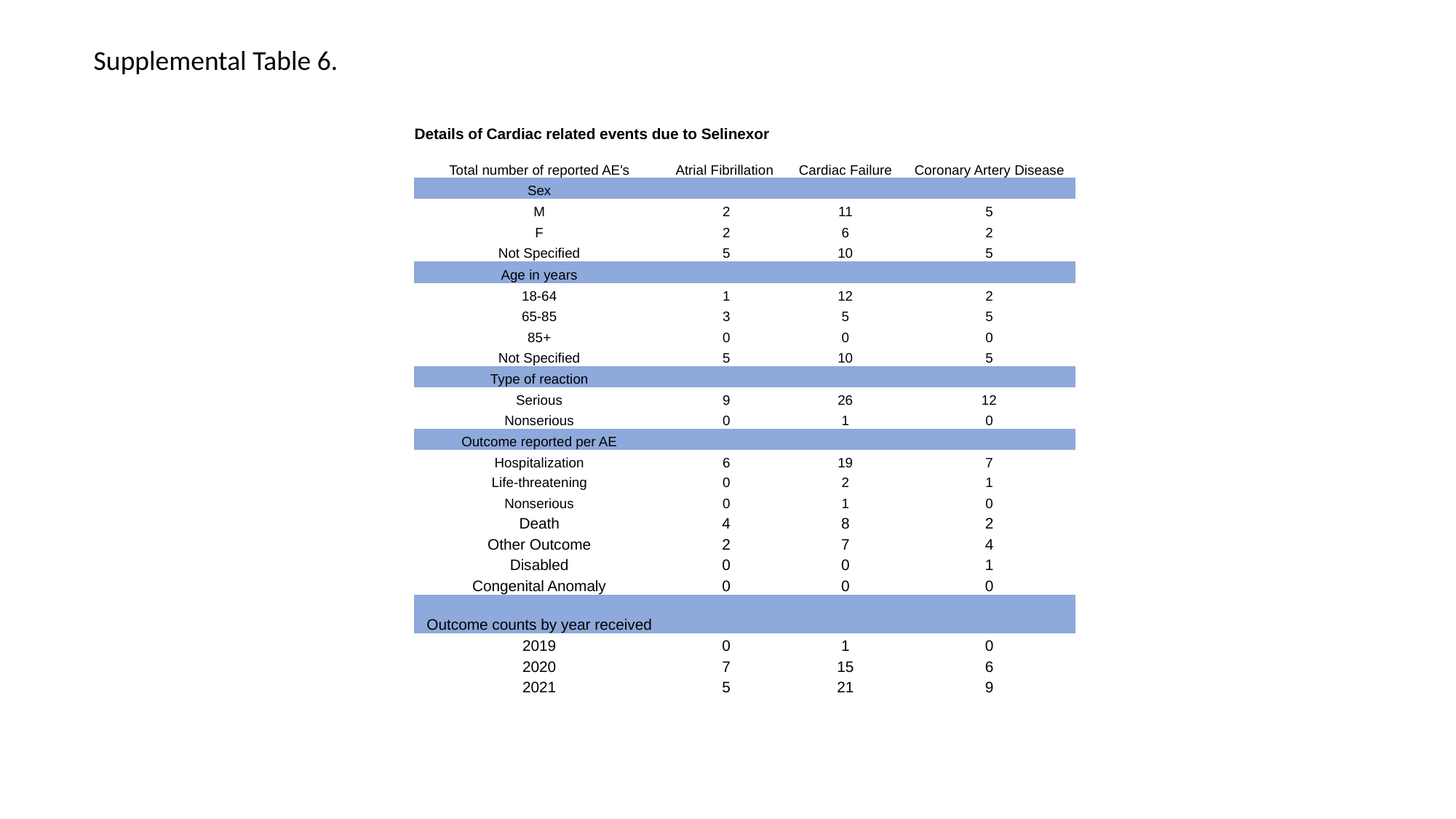

Supplemental Table 6.
| Details of Cardiac related events due to Selinexor | | | |
| --- | --- | --- | --- |
| Total number of reported AE's | Atrial Fibrillation | Cardiac Failure | Coronary Artery Disease |
| Sex | | | |
| M | 2 | 11 | 5 |
| F | 2 | 6 | 2 |
| Not Specified | 5 | 10 | 5 |
| Age in years | | | |
| 18-64 | 1 | 12 | 2 |
| 65-85 | 3 | 5 | 5 |
| 85+ | 0 | 0 | 0 |
| Not Specified | 5 | 10 | 5 |
| Type of reaction | | | |
| Serious | 9 | 26 | 12 |
| Nonserious | 0 | 1 | 0 |
| Outcome reported per AE | | | |
| Hospitalization | 6 | 19 | 7 |
| Life-threatening | 0 | 2 | 1 |
| Nonserious | 0 | 1 | 0 |
| Death | 4 | 8 | 2 |
| Other Outcome | 2 | 7 | 4 |
| Disabled | 0 | 0 | 1 |
| Congenital Anomaly | 0 | 0 | 0 |
| Outcome counts by year received | | | |
| 2019 | 0 | 1 | 0 |
| 2020 | 7 | 15 | 6 |
| 2021 | 5 | 21 | 9 |
